# The genetic architecture of obsessive-compulsive disorder: alleles across the frequency spectrum contribute liability to OCD

**DOI:** 10.1101/2021.01.26.21250409

**Authors:** Behrang Mahjani, Lambertus Klei, Manuel Mattheisen, Matthew W. Halvorsen, Abraham Reichenberg, Kathryn Roeder, Nancy L. Pedersen, Julia Boberg, Elles de Schipper, Cynthia M. Bulik, Mikael Landén, Bengt Fundín, David Mataix-Cols, Sven Sandin, Christina M. Hultman, James J. Crowley, Joseph D. Buxbaum, Christian Rück, Bernie Devlin, Dorothy E. Grice

**Affiliations:** Seaver Autism Center for Research and Treatment, Icahn School of Medicine at Mount Sinai, New York, NY, USA; Division of Tics, Obsessive-Compulsive Disorder (OCD) and Related Disorders, Icahn School of Medicine at Mount Sinai, New York, NY, USA; Department of Psychiatry, Icahn School of Medicine at Mount Sinai, New York, NY, USA; Department of Medical Epidemiology and Biostatistics, Karolinska Institutet, Stockholm, Sweden; Department of Psychiatry, University of Pittsburgh School of Medicine, Pittsburgh, Pennsylvania, USA; Department of Psychiatry, Dalhousie University, Halifax, Nova Scotia, Canada; Department of Biomedicine, Aarhus University, Aarhus, Denmark; Department of Genetics, University of North Carolina at Chapel Hill, North Carolina, USA; The Mindich Child Health and Development Institute, Icahn School of Medicine at Mount Sinai, New York, NY, USA; Department of Statistics, Carnegie Mellon University, Pittsburgh, Pennsylvania, USA; Computational Biology Department, Carnegie Mellon University, Pittsburgh, Pennsylvania, USA; Centre for Psychiatry Research, Department of Clinical Neuroscience, Karolinska Institutet, Stockholm, Sweden; Department of Psychiatry, University of North Carolina at Chapel Hill, Chapel Hill, North Carolina, USA; Department of Nutrition, University of North Carolina at Chapel Hill, Chapel Hill, North Carolina, USA; Institute of Neuroscience and Physiology, University of Gothenburg, Gothenburg, Sweden; Department of Genetics and Genomic Sciences, Icahn School of Medicine at Mount Sinai, New York, NY, USA; Friedman Brain Institute, Icahn School of Medicine at Mount Sinai, New York, NY, USA; Department of Neuroscience, Icahn School of Medicine at Mount Sinai, New York, NY, USA; Health Care Services, Region Stockholm, Stockholm, Sweden

**Author notes:** contributed equaliy (Behrang Mahjani, Lambertus Klei, Manuel Mattheisen, Christian Rück, Bernie Devlin, Dorothy E. Grice). **Location of work and address for reprints**: Dorothy E. Grice, M.D., 1425 Madison Avenue, New York, NY 10029. **Disclosures**: Authors report no financial relationships with commercial interests.

## Abstract

**Objective:** Obsessive-compulsive disorder (OCD) is known to be substantially heritable; however, the contribution of common genetic variation across the allele frequency spectrum to this heritability remains uncertain. We use two new, homogenous cohorts to estimate heritability of OCD from common genetic variation and contrast results with prior studies.

**Methods:** The sample consisted of 2096 Swedish-born individuals diagnosed with OCD and 4609 controls, all genotyped for common genetic variants, specifically >400,000 single nucleotide polymorphisms (SNPs) with minor allele frequency (MAF) ≥ 0.01. Using genotypes of these SNPs to estimate distant familial relationships among individuals, we estimated heritability of OCD, both overall and partitioned according to MAF bins.

**Results:** We estimated narrow-sense heritability of 28% (SE=4%). The estimate was robust, varying only modestly under different models. Contrary to an earlier study, however, SNPs with MAF between 0.01 and 0.05 accounted for 8% of heritability and estimated heritability per bin roughly follows expectations based on a simple model for SNP-based heritability.

**Conclusions:** These results indicate that common inherited risk variation (MAF ≥ 0.01) accounts for most of the heritable variation in OCD. SNPs with low MAF contribute meaningfully to the heritability of OCD and the results are consistent with expectation under the “infinitesimal model,” where risk is influenced by a large number of loci across the genome and across MAF bins.

## 1. Introduction

Obsessive-compulsive disorder (OCD) is a serious and often long-lasting psychiatric disorder characterized by intrusive and unwanted thoughts, images, or urges (obsessions) that are typically linked to ritualized behaviors (compulsions) (1–4). OCD affects 1-3% of the population and multiple studies provide reliable evidence for a significant genetic contribution to risk (1, 3–6), as well as a role for environmental factors impacting risk (7, 8). The heritability of OCD, historically estimated by analysis of twin and family studies and within the context of the ACE model (additive genetic, also known as narrow sense heritability, A; shared environment, C; and nonshared environment, E), is reported to be 35-50% (1, 4, 8–13).

As an alternative to the analysis of recurrence risk for OCD within pedigrees, heritability can also be estimated from individuals drawn from a population who have no obvious familial relationships, as long as they have been characterized for genetic variation across their genomes. Usually, this genetic characterization employs genotypes of single nucleotide polymorphisms (SNPs) for which alleles are common in the population. In this approach, which we will call SNP-based, the central idea is that the multiplicity of SNP genotypes allows estimation of familial relationships, albeit distant, among subjects as well as the covariance of their phenotypes, and these are the key elements for estimating heritability. When the heritability of OCD is computed in this manner, estimates range from 25-43% (5, 14, 15).

It is useful to compare the heritability results from family-based and SNP-based approaches. Family-based studies, being more direct, typically yield estimates of heritability with lower standard errors, whereas the inaccuracy of estimating distant relationships from genetic data tends to produce fuzzier estimates. Family-based estimates also tend to yield higher estimates of heritability because the familial covariance traces to both rare and common genetic variation, whereas SNP-based estimates mostly arise from covariance due to common genetic variants. Looking at the results summarized above, one might conclude that this is also operating for OCD, i.e., that family-based studies are producing higher heritability estimates than SNP-based studies.

However, in an influential paper by Davis and colleagues (5), there was no evidence for heritability from SNPs with minor allele frequency (MAF) < 0.05 and over 60% of total heritability mapped to the most common variants (MAF > 0.3). If this observation were true, it could have profound implications for which evolutionary forces shaped this unusual mapping of risk alleles to their population frequency distribution. For example, balancing selection, where multiple alleles are maintained in the gene pool of a population at frequencies larger than expected from genetic drift alone may play a role in OCD.

At the same time, other studies have implicated rare variants in risk for OCD (16–19). Thus, the contribution of inherited genetic variation across the allelic frequency spectrum to the risk of OCD remains uncertain and worthy of further study, as it impacts both our understanding of processes underlying OCD risk architecture and rational study design. Here, using a substantially larger sample compared to previous studies and new genetic data from the Swedish population, we estimate SNP-based heritability for OCD.

## 2. Methods

### 2.1 Study population

Ethical approvals were obtained from the Institutional Review Board (IRB) at the Icahn School of Medicine at Mount Sinai, New York, NY, and the Regional Ethical Review Board in Stockholm. We used Swedish OCD cases collected through two studies: the EGOS cohort (Epidemiology and Genetics of Obsessive-compulsive disorder and chronic tic disorders in Sweden) (20) and the NORDiC cohort (Nordic OCD and Related Disorders Consortium) (21).

In the EGOS cohort, individuals born between 1954 and 1998, with at least two clinical diagnoses of OCD in the Swedish National Patient Register (NPR), were eligible for inclusion (20). In the Swedish site of the NORDiC cohort, individuals with OCD were recruited from specialty OCD and related disorder clinics across Sweden (21). Genotype data on the global screen array (GSA) were collected for 1108 individuals from the EGOS cohort and 1107 individuals from the NORDiC cohort.

A sample of 4738 controls from the LifeGene cohort was available for this study. LifeGene is a prospective population-based cohort of around 50,000 individuals in Sweden (22). The samples were available in four batches: LifeGene-EGOS (n=1444), LifeGene-NORDiC (n=500), LifeGene-ANGI-Wave-1 (n=1500), and LifeGene-ANGI-Wave-2 (n=1500). LifeGene-ANGI controls were previously used in a study of anorexia nervosa (AN) (23); they were mostly females (2935 females and 65 males), and all individuals with a diagnosis of AN were previously removed from this batch. All controls were genotyped using GSA.

### 2.2 Quality control

All OCD cases, LifeGene-EGOS controls, and LifeGene-NORDiC controls were genotyped in the same laboratory but in different batches. GenomeStudio’s genotyping module was used to re-call genotypes on the joint data.

Quality control (QC) was first carried out on three batches of samples that may differ in key variables: 1) all cases, LifeGene-EGOS controls, and LifeGene-NORDiC, 2) LifeGene-ANGI-Wave-1 controls, and 3) LifeGene-ANGI-Wave-2 controls. We employed the following QC steps using PLINK 2.0 (Supplementary Materials Tables S1-S3): individuals were removed who had a genotype non-call rate > 0.05, were discrepant for nominal versus genetically-determined sex, or had low heterozygosity (< -3SD from the mean); a SNP was removed if its non-call rate for genotypes was > 0.05, its MAF < 0.01, or it had Hardy–Weinberg equilibrium (HW) p-value < 0.00125. Gemtools was used to choose individuals with European ancestry where indicated (Supplementary Materials Figure S1).

We next used the McCarthy tool to match the SNPs to 1000 Genomes, and Genotype Harmonizer software (automatic strand alignment software) to align the different cohorts (24). After QC, we merged the cohorts based on the set of all intersecting SNPs and performed additional QC as noted in Supplementary Materials. The final data set included 2096 cases and 4609 controls, with 405,105 SNPs (Table 2).

**Table 1.** Characteristics of the cohorts.

| Characteristics | Category | EGOS | NORDiC |
| --- | --- | --- | --- |
| Total number of OCD cases |  | 1108 | 1107 |
| Sex, count (%) | Females | 692 (63%) | 651 (59%) |
| Comorbidities, count (%) | CTD (ICD-10: F95) | 100 (9%) | 43 (5%)<sup>1</sup> |
|  | ADHD (ICD-10: F90) | 40 (4%) | 83 (10%)<sup>1</sup> |
|  | Bipolar Disorder (ICD-10: F31) | 33 (3%) | 93 (11%)<sup>1</sup> |
|  | Phobic anxiety disorders (ICD-10: F40) | 19 (2%) | 98 (12%)<sup>1</sup> |
|  | Other anxiety disorders (ICD-10: F41) | 112 (10%) | 106 (13%)<sup>1</sup> |
|  | Autistic disorder (ICD-10: F84.0) | 3 (0.3%) | 8 (1%)<sup>1</sup> |
|  | Asperger's syndrome (ICD-10: F84.5) | 40 (4%) | 51 (6%)<sup>1</sup> |
|  | Intellectual disability (ICD-10: F71-73) | 7 (0.6%) | 2 (0.2%)<sup>1</sup> |
|  | At least one psychiatric diagnosis | 417 (37%) | 424 (53%)<sup>1</sup> |
| Diagnosis age/Age at first symptom (p5, Median, p95)<sup>2</sup> |  | (12,21,34) | (5,12,30)<sup>3</sup> |

<sup>1</sup>283 individuals have missing values (n=804). <sup>2</sup>p5 and p95 are the 5th and 95th percentiles. <sup>3</sup>437 missing values (n=670). OCD: obsessive-compulsive disorder, CTD: chronic tic disorders, ADHD: attention-deficit/hyperactivity disorder.

**Table 2.**
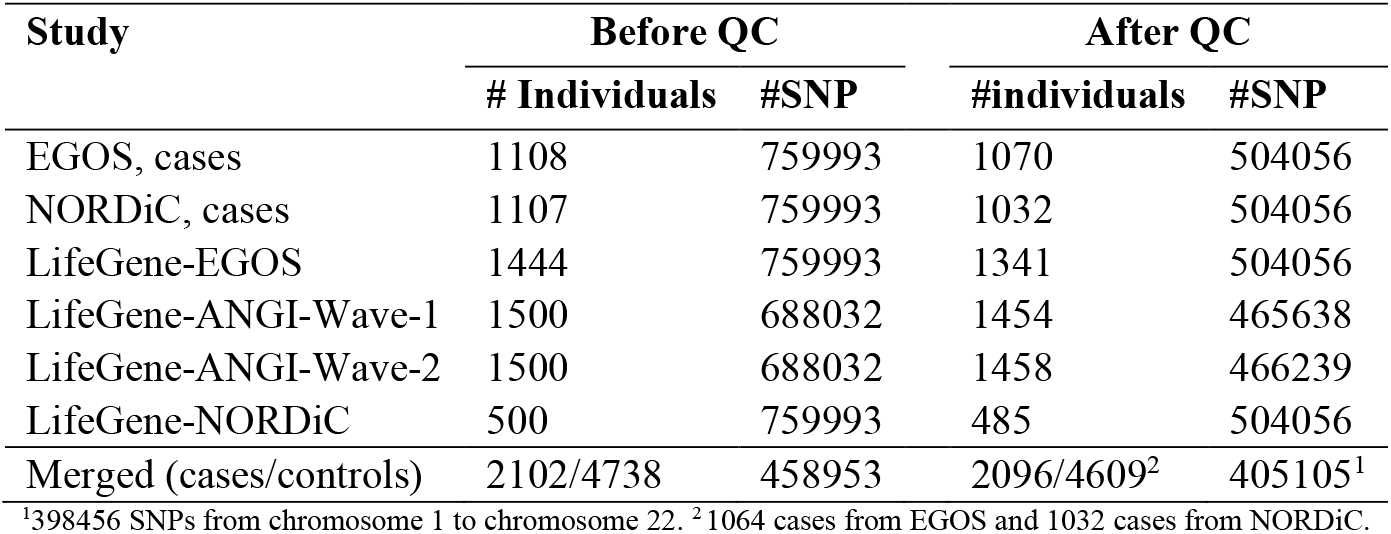
Summary of data before and after quality control.

### 2.3 Statistical analysis

We used the Genome-wide Complex Trait Analysis (GCTA) program version 1.26.0 to estimate the genetic relationship matrix (GRM) between all pairs of individuals from SNPs (25). Then, we used PLINK 2.0 to extract the top principal components (PCAs) from the variance-standardized relationship matrix (for more details, see Supplementary Materials). We performed restricted maximum likelihood (REML) analysis, implemented in GCTA, to estimate heritability of OCD attributable to SNP genotypes. Because the OCD diagnosis is dichotomous, we scaled the phenotypic variance to an underlying liability scale using the population prevalence of 1%, similar to our most recent estimate of population prevalence in Sweden using data from the Swedish national registers (1) (for more details, see Supplementary Materials, where we also provide results for 2% prevalence).

To evaluate the sensitivity of estimates of SNP-based heritability to modeling approaches, we assessed the data in multiple ways: **(1)** Included all affected and unaffected individuals born in Sweden, of whom most, but not all, were of Swedish/European genetic ancestry; use all 405,105 high quality, genotyped SNPs for analysis. The sampling in **(1)** is consistent with our previously-published, family-based analyses and will be our primary analytical approach. **(2)** Pruned SNPs according to linkage disequilibrium (LD) to obtain a smaller set of 185,066 largely independent SNPs. **(3)** Limit the sample to individuals of European genetic ancestry. **(4)** Removed all individuals for whom there is also a fifth degree or greater relative in the sample. **(5)** Analyzed only pairs of affected and unaffected individuals, matched on two dimensions of genetic ancestry using the function *pairmatch* in the package *optmatch* in R (1-to-1 fullmatch) (Supplementary Materials). **(6)** As in **(5)**, using only individuals of European ancestry. Pair matching, as done in **(5)** and **(6)**, is a common epidemiological approach for controlling confounding (here, differences in ancestry in cases versus controls) and has been shown to be useful for genetic studies (26–28). Note **(3)-(6)** use all high-quality SNPs.

We also estimated heritability partitioned by chromosomes and MAF bins and compared the results with those from Davis *et al*. (5). Following Davis, we created six MAF bins: 0.01-0.05, 0.05-0.1, 0.1-0.2, 0.2-0.3, 0.3-0.4, and 0.4-0.5. For each bin, we computed a GRM, estimated heritability, and then combined them (using --mgrm in GCTA). This allows for the effects of LD to be partitioned by the REML.

## 3. Results

Our study population included 2096 cases and 4609 controls after quality control was completed. There were 61% females among our cases. Based on Principal Component Analysis (PCA; Figure S2), we used the first six PCAs as covariates to adjust for variation in ancestry in all heritability analyses. As a check for compatibility of cohorts, we first estimated heritability by treating EGOS and NORDiC controls as cases and LifeGene-ANGI controls as controls. Heritability was estimated at 0.0001% (SE = 5%). These results show that the control cohorts were homogeneous. Next, we estimated heritability of OCD for the full sample, contrasting OCD cases to controls and yielding an estimate of 28% (SE=4%) for a population prevalence of 1%.

Technically, heritability is first estimated on the observed scale, namely dichotomous OCD diagnosis; however, heritability on the continuous liability scale is more interpretable and so is usually reported. Heritability can be transformed from the observed to the continuous liability scales because they are functions of prevalence (29). To determine how sensitive our heritability estimate was to prevalence, we varied it between 0.5%-3% and found heritability to vary between 24%-37% (Supplementary Materials Table S4).

We next performed a set of sensitivity analyses by different treatments of the data, as described in Methods, and found the estimates to be quite robust (Table 3). Notably, although analyses suggested that EGOS and NORDiC cases had slightly different ancestry distributions, the results in Table 3 show that our adjustments for ancestry were sufficient to compensate for these differences (Figures S4-S9, Table S5). In addition, we did not observe a significant difference in heritability between the EGOS and NORDiC cases (Table S5).

**Table 3.** Estimates of heritability of OCD under various treatments of the data.

|  | #Cases/Controls | Heritability (SE) |
| --- | --- | --- |
| All cases and controls | 2096/4609 | 28% (4%) |
| All cases and controls (using 185066 SNPs after LD pruning) | 2096/4609 | 29% (4%) |
| Individuals with European ancestry only | 1758/3707 | 29% (5%) |
| All third cousins or closer relatives removed | 1816/3925 | 30% (4%) |
| Ancestry-matched (1-to-1 fullmatch) cases and controls | 2096/2096 | 23% (6%) |
| Ancestry-matched (1-to-1 fullmatch) cases and controls, European ancestry | 1758/1758 | 26% (7%) |

### 3.1 Heritability analysis partitioned by MAF bins

Having established that a substantial portion of OCD traces to common variation, we next addressed an important issue about its nature. Specifically, in an earlier study, Davis *et al*. (5) found that alleles with MAF < 0.05 did not contribute meaningfully to the heritability of OCD (0.0001% of total heritability). To compare our results to those in Davis *et al*. (5), we estimated the portion of total heritability for groups of autosomal SNPs with distinct allele frequencies, grouping the SNPs into six bins based on their MAF (Figure 1; Table S10): 0.01-0.05, 0.05-0.1, 0.1-0.2, 0.2-0.3, 0.3-0.4, and 0.4-0.5. For all the bins, we included the first six PCAs as covariates and set population prevalence to 0.01. Estimates of the portion of total heritability for the bins were distributed differently between these two studies (Figure 1; Table S10). Curiously, although the total heritability of the first two bins (MAF < 0.1) was similar across studies, 2.5% for our study versus 4% for Davis, estimates for specific bins were not similar; in the Davis *et al*. study, the MAF bin from 0.01-0.05 accounted for essentially no heritability (0.0001%) whereas our estimate was much larger (2.5%) (Figure 1; Table S10). A portion of the difference could be due to the number and nature of the SNPs falling in this bin: there were approximately ten times more genotyped SNPs falling into this bin in the current study compared with Davis *et al*. (Table S10); however, Davis also imputed genotypes for over 2 million SNPs for this bin and those genotypes did not alter their heritability estimate in that bin (see their Table 2).

**Figure 1.**
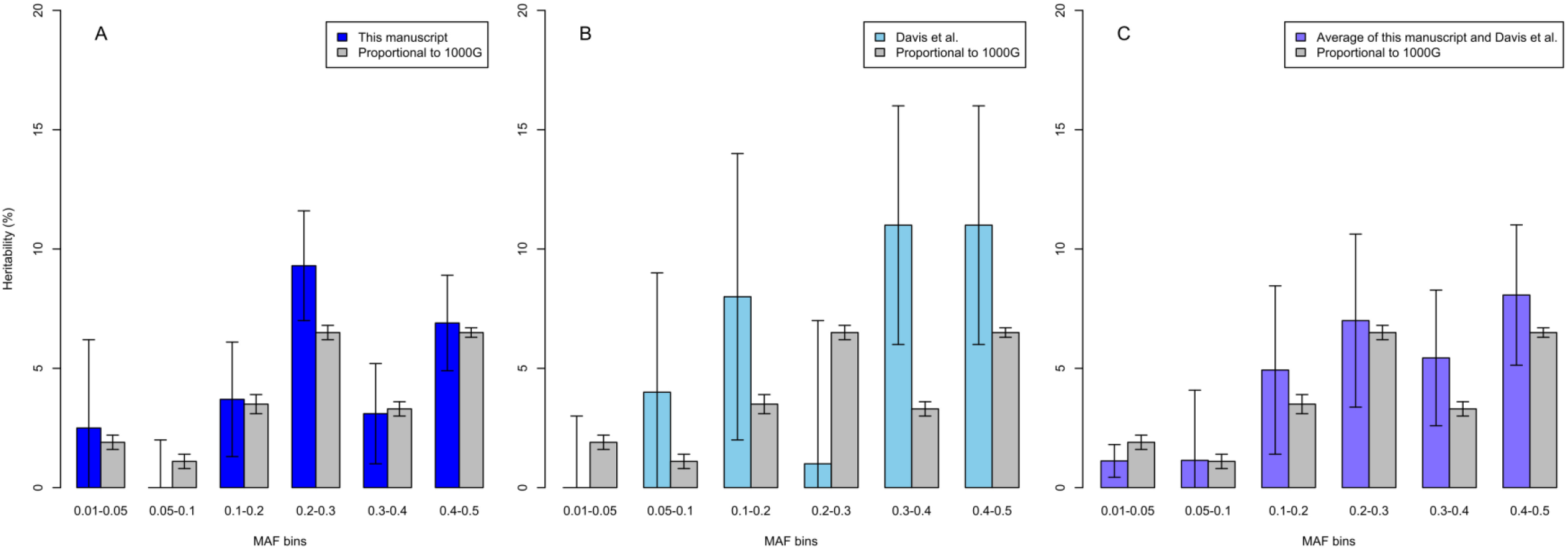
Estimates of heritability partitioned by MAF bins from the results in A) this study, B) Davis *et al*. (5) and, C) weighted averages (weights proportional to the inverse of variance) of this study and Davis *et al*. In each panel, we also show the estimate of heritability for each bin from 1000G data, presented as the mean of heritability for that bin for ten samples of size 108K SNP, where sampling from each bin was proportional to the percentage of SNPs in that bin from 1000G data. Note that the SE for this latter analysis is the standard error of the sample mean for the ten samples and is not directly comparable to the SNP-based SE. Correlations with 1000G data were 0.96, p-value=0.002, for panel A; 0.25, p-value=0.64, for panel B; and 0.94, p-value=0.005, for panel C.

To investigate these differences, we estimated what the expected portion of total heritability in these bins should be. First, we observed that the percentages of the total SNPs in each bin were distributed differently in comparison to 1000 Genomes data (for SNPs with MAF > 0.01) (Tables S6 and S7), which we would expect is more representative of variation in the general population. For example, 45.6% of the SNPs in our study had MAF between 0.01 and 0.05, while 29.5% of SNPs in 1000 Genomes data had MAF between 0.01 and 0.05. Under the standard quantitative genetic “infinitesimal model” (also referred to as the “polygenic model”), it is reasonable to assume the effect of all risk SNPs is equal. With this assumption, we then explored various models to predict the expected heritability in each MAF bin (Figure 1; Tables S6-S8; Figures S10 and S11).

The model that best fit the data was one in which risk alleles were sampled proportional to their occurrence in 1000 Genomes data, with a goodness-of-fit R^2^ = 0.56. Notably, the largest proportion of expected heritability was not explained by SNPs in the higher frequency allele bins (0.3-0.4 and 0.4-0.5), contrary to what was observed in Davis *et al*. (5). In addition, we observed that SNPs with low MAF (0.01-0.05) are expected to account for 8.3% of the heritability under this model, similar to the 9.7% that we observed and in contrast to Davis where low MAF SNPs accounted for almost no heritability. These discrepancies and the smaller ones observed in our study track with sample size: the sample size for Davis *et al*. (1061 cases and 4236 controls) was smaller than our current study and variance of estimates are a direct function of sample size. The Davis *et al*. sample (5) could also be more ancestrally heterogeneous than our present Swedish sample, and this could play some role in increasing variance in their estimates. Combining results from both studies demonstrated strong concordance with expectation (Figure 1C).

### 3.2 Heritability analysis partitioned by chromosomes

Under the infinitesimal model, SNPs affecting heritability of OCD (or any trait) should be scattered randomly across chromosomes, so that heritability per chromosome should track with chromosome length. This is observed in our study (Figure 2) and there is a significant correlation between heritability per chromosome and length (r = 0.64, p-value = 0.001). Chromosome 13 had the lowest heritability, significantly lower than what would be expected under the uniform distribution model.

**Figure 2.**
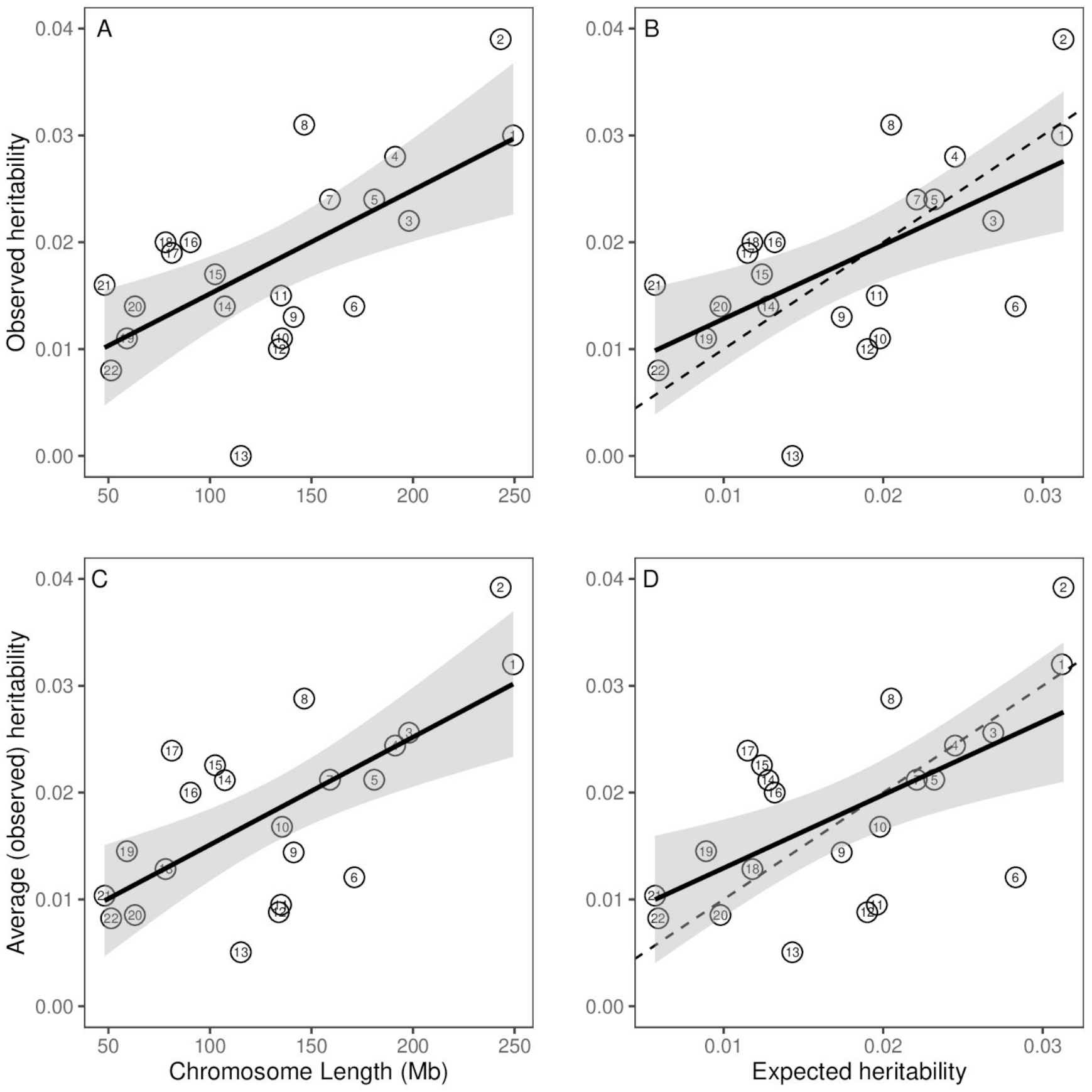
Estimates of heritability partitioned by chromosome. A) The observed heritability by chromosome length and the 95% confidence interval (CI) for the regressed line (R^2^=0.39, p-value=0.001); B) The observed heritability by expected heritability and the 95% CI for the regressed line (R^2^=0.34, p-value=0.003); C) The weighted average observed heritability by chromosome length and the 95% CI for the regressed line (average over this manuscript and Davis *et al*. study) (R^2^=0.42, p-value=0.001), the results for chromosome 21 and 22 are overlapping; and D) The weighted average heritability by expected heritability and the 95% CI for the regressed line (R^2^=0.34, p-value=0.028), the results for chromosome 21 and 22 are overlapping. The dashed lines have slope one and intercept zero (observed=expected).

As noted above, the noisy nature of these results can likely be attributed to relatively small sample size for this type of analysis. We conjectured that if this were the case, and assuming both study samples were homogeneous, combining the Davis *et al*. heritability estimates and our heritability estimates, per chromosome, would produce a somewhat better fit between heritability per chromosomes and length. This result is confirmed in Figure 2C-1D; the fit of the regression for this weighted average heritability (weights proportional to the inverse of variance), R^2^ = 0.42, is better than the fit for our sample alone, R^2^=0.39. Furthermore, note that chromosome 6, which had very low heritability in Davis *et al*., shows reasonable heritability in both our analyses and in the combined data, again suggesting small sample sizes are driving some of the results.

## 4. Discussion

Common genetic variation – variants shared among many individuals in a population and most frequently SNPs – has been found to play a role in liability for most psychiatric disorders, including OCD. Open questions remain about the impact on risk due to common variation, including how much of the heritability of OCD it accounts for and how it is partitioned across the frequency spectrum of alleles. These are important questions for a variety of reasons. For example, both schizophrenia and autism spectrum disorder demonstrate high heritability (30, 31) and much of it traces to common genetic variation. Yet rare variation with a damaging impact on gene function, especially *de novo* variation, plays a larger role in overall autism spectrum disorder risk than in overall schizophrenia risk (30, 32, 33); e.g., in Singh *et al*. (33), *de novo* protein truncating variants were found to be fourfold more common in individuals with autism than schizophrenia when they evaluated evolutionarily-constrained genes. This difference is critical for clinical genetics, genetic counseling, and possibly treatment. It also could be relevant for disentangling evolutionary processes underlying different psychiatric disorders, consistent with stronger natural selection on autism than schizophrenia. Finally, it would impact study design (if, for example, rare variants contribute little to OCD heritability).

Here we evaluate whether a substantial portion of the heritability of OCD traces to common variation, as it does for autism and schizophrenia, and characterize its frequency spectrum, which is directly relevant to evolutionary processes. For example, in an early study estimating heritability of OCD from common variation, results in Davis *et al*. (5) suggested that alleles with the highest frequencies, i.e., those with MAF>0.3, account for the bulk of SNP-based heritability of OCD. Such a strong pattern would suggest that OCD was under strong balancing selection. By sampling individuals with OCD from the Swedish population, as well as a larger sample of unaffected (control) individuals, we were able to address these questions. Our analyses of over 2000 individuals diagnosed with OCD and twofold more unaffected individuals, each genotyped across their genome via >400,000 SNPs, yielded an OCD heritability estimate of 28% (SE=4%), a robust estimate (Table 3).

Moreover, when we assumed SNPs contributed equally to risk for OCD, regardless of MAF, we obtained good fit between estimated OCD heritabilities from MAF bins of our sample and what was expected based on the distribution of MAF in 1000 Genomes data (Figure 1). SNPs affecting risk appear to be distributed at random over chromosomes because size was a good predictor of a chromosome’s contribution to total heritability (Figure 2). Chromosome 13 showed the poorest fit to this model, which may be partially explained by it having one of the lowest gene densities (6.5 genes per Mb) among human chromosomes. All of these results fit expectations of the infinitesimal quantitative genetics model.

In terms of estimated heritability from common variation, our results compare favorably with previous studies of OCD. Published estimates of SNP-based heritability, based on different samples from different populations, range from 25-43% (5, 14, 15). Thus, all studies have converged on a substantial contribution of common variation to the heritability of OCD, showing notable consistency. There are some differences, however. Notably, the recent study by Davis and colleagues suggest that only SNPs with substantial frequency in their population sample (MAF > 0.05) contribute to this heritability and the contribution to heritability tends to increase with increasing MAF.

In light of our findings, we found their results intriguing: an increasing heritability associated with MAF is appealing because the contribution to heritability of any SNP of frequency *p* is *2p(1-p)a*^2^, where the SNP’s effect *a* can be assumed to be roughly equal over all SNPs under the infinitesimal model; on the other hand, it seems unlikely that low MAF SNPs have no contribution to heritability because there are so many of them in the human genome (Table S10). Our results from Sweden argue that these low MAF SNPs do contribute to OCD heritability, their contribution is roughly in proportion to the frequency spectrum of alleles, and can be assumed to be of similar effect (i.e., *a*) across the frequency spectrum. Thus, our results show that future studies of less common and even rare alleles are also informative for OCD etiology, with the caveat that effects of risk alleles of very low frequency can be difficult to detect by case-control methods.

Another interesting contrast is the evidence for heritability across chromosomes. Davis *et al*. observed essentially no heritability for OCD on chromosome 6, which encodes both the HLA and histone gene clusters, and extremely high heritability on chromosome 15. In discussing these results, the authors suggest that chromosome 15 has an outsized contribution to OCD risk and that the HLA locus is effectively excluded from OCD risk. Given the contrasting results in our study, and in our analyses combining results from both studies, we again conclude that the data are consistent with the infinitesimal model and that smaller sample sizes might account for results that diverge from expectation.

The previous work by Davis and colleagues involved about 50% fewer OCD cases and the variance in any estimate is a direct function of sample size. It is also possible that the Davis study had a different distribution of distantly related individuals than our relatively homogeneous sample from Sweden. Accuracy of SNP-based heritability diminishes as the fraction of very distantly related pairs, relative to all relative pairs, increases. Consistent with estimates from both studies being noisy, when we combined the Davis *et al*. results to obtain new estimates of average heritability per allele bin and heritability per chromosome, the average fit expectation was better than in either study alone.

The present study had strengths and limitations. We used OCD cases from the EGOS and NORDiC cohorts, the two largest OCD studies in Sweden to examine the role of genetic and environmental factors. The EGOS cohort utilized the NPR for its sampling frame, thus it is an epidemiological cohort minimizing selection biases, while the NORDiC recruited through specialty OCD clinics across Sweden, a sampling frame more typical of case-control studies. This difference in sampling frames could introduce heterogeneity into our study. Nonetheless, when we evaluated this possibility by estimating the heritability induced by contrasting OCD cases from EGOS to OCD cases from NORDiC, and doing the same for controls, both estimated heritabilities were not significantly different from zero. Hence, while there could be subtle heterogeneity between the cohorts, it must be small. Furthermore, for both cohorts, reliance on inclusion as a result of individuals seeking care at mental health hospitals/clinics can inadvertently exclude those with milder forms of the disorder who may seek treatment from primary care providers and/or those who do not present to clinical services at all. If such individuals were included and if their genetic architecture were different from our current OCD case sample, it would impact the estimated heritability. By restricting cases to individuals in Sweden, we had a genetically homogeneous sample, which minimized the risk of confounding due to population stratification and facilitated the combining of the cohorts. Nonetheless, it does limit the generalizability of our results. However, after combining our results with those of Davis *et al*., we observe results that fit expectation, thus suggesting that the results are likely to relevant for most populations.

Prior to the advent of dense genotyping, the heritability of a trait was typically estimated from its distribution within pedigrees. These kinds of studies continue to this day, in large part because they capture heritability due to both common and rare inherited genetic variation. It is thus interesting to compare our SNP-based heritability estimate from common variation, 28%, to that from Swedish families, 35-50% (1, 4). This comparison suggests that while the majority of inherited liability for OCD in Sweden traces to common genetic variation, rare variation contributes to OCD liability as well, but to a lesser degree, consistent with the findings to date regarding rare variation and risk for OCD (16–19).

In summary, our results demonstrate that the majority of inherited liability for OCD in Sweden traces to common genetic variation. Moreover, our results show that the distribution of risk as a function of allele frequency is consistent with expectations, indicating that balancing selection, or other more complex evolutionary forces, are not strongly at play in OCD. Furthermore, our results indicate that risk for OCD is distributed across the genome as expected and that results presented here and in prior studies are consistent with the infinitesimal model for OCD. Finally, our results support the continued study of rare variation, both inherited and de novo, in OCD risk.

## Data Availability

The data that support the findings of this study are available on request from the corresponding author, [DEG]. The data are not publicly available.

## Acknowledgments

This study was supported by a grant from the Friedman Brain Institute (DEG), and the Beatrice and Samuel A. Seaver Foundation (DEG, SS, JDB, BM); the Mindworks Charitable Lead Trust (DEG); the Stanley Center for Psychiatric Research (DEG and JDB); and NIMH grant R01MH124679 (DEG), R37MH057881 (BD/KR) and R01MH110427 (JC); the Swedish Research Council (grant numbers 2015-02271, 2018-02487) (DM-C and CR), CIMED and Region Stockholm (CR).

Funding for some of the control samples was from the Anorexia Nervosa Genetics Initiative (ANGI), an initiative of the Klarman Family Foundation (PI: Bulik). Dr. Bulik is also supported by (R01MH120170, R01MH119084 PI: Bulik; U01 MH109528 PI: Sullivan); the Swedish Research Council (Vetenskapsrådet, award: 538-2013-8864); Brain and Behavior Research Foundation Distinguished Investigator Grant; and Lundbeck Foundation (Grant no. R276-2018-4581).

The computation was performed on resources provided by SNIC through Uppsala Multidisciplinary Center for Advanced Computational Science (UPPMAX) under Project sens2018605.

LifeGene was supported by the Torsten and Ragnar Söderbergs Foundation, Karolinska Institutet, Stockholm County Council, and AFA Insurance. LifeGene is a core facility at Karolinska Institutet.

## Author Contributions

Study concept and design: JDB, BD, DEG, LK, BM

Acquisition, analysis, or interpretation of data: JDB, JB, JC, BD, DEG, MH, LK, BM, MM, EdS, DMC, CR, NLP

Drafting of the manuscript: JDB, BD, DEG, LK, BM

Critical revision of the manuscript for important intellectual content: All authors.

Statistical analysis: BD, LK, BM

Obtained funding: JDB, JC, DEG, MC, MM, DR, DMC, CR

Study supervision: JDB, BD, DEG

