## Supplemental material for "The genetic architecture of obsessive-compulsive disorder: alleles across the frequency spectrum contribute liability to OCD"

### Supplemental Information

#### Quality control

After quality control described in the body of the manuscript, we merged the cohorts and performed the following additional quality control steps:

- removed 135 subjects, 6 diagnosed with OCD, deemed to be close relatives (pihat > 0.2).

By contrasting allele frequencies in the different cohorts using measure of allelic variation such as Fst, and by analyzing only individuals genetically identified as of European ancestry, we removed variants with:

- fixation index (FSt) > 0.005 (168 variants) between controls,
- FSt > 0.005 (6 variants) between all cohorts,
- FSt > 0.005 (4 variants) between EGOS and controls,
- FSt > 0.005 (2 variants) between NORDiC and controls,
- missingness in a cohort > 0.02 (11390 variants),
- (max – min) allele frequency across the control > 0.03 (39335 variants).

Next, we sought to remove poorly called SNPs by contrasting allele frequencies from LifeGene (iCON and NORDiC) controls versus LifeGene-ANGI controls using a standard logistic association test, as would be used for a GWAS. We removed 189 variants with p-value < 1e-4.

We removed SNPs with a significant difference in missingness between OCD cases and controls |(missingness – mean miss missingness)| >0.01 (2754 variants).

The final dataset had 2096 cases and 4609 controls, with 405105 SNPs (53848 variants were removed after merging the cohorts).

**Table S1.** Details of QC for EGOS cases, NORDiC cases, LifeGene iCON, LifeGene NORDiC.

| | Individuals | SNPs | Removed individuals in each step | Removed SNPs in each step |
|---|---|---|---|---|
| cases/controls | 2215/1943 | 759993 | - | - |
| **Phase 1: Pre-QC** | | | | |
| a. Check duplicate marker names | 2215/1943 | 759993 | - | 0 |
| b. SNPs not containing rs as part of the name | 2215/1943 | 708521 | - | 51472 |
| c. Remove SNPs without location | 2215/1943 | 701511 | - | 7010 |
| d. Remove SNPs on PAR and MT | 2215/1943 | 699608 | - | PAR:927, MT:976 |
| e. Remove all homozygous SNPs | 2215/1943 | 696155 | - | 3453 |
| f. INDELs | 2215/1943 | 687102 | - | 9053 |
| g. Remove SNPs sharing the same location | 2215/1943 | 687102 | - | 0 |
| h. Remove ambiguous SNPs | 2215/1943 | 677246 | - | 9856 |
| i. Non call rate on SNPs (0.15) | 2215/1943 | 675308 | - | 1938 |
| **Phase 2: QC on individuals** | | | | |
| a. Check for duplicate samples IDs | 2215/1943 | 675308 | 0 | - |
| b. Remove samples with plating issues | 2215/1943 | 675308 | 0 | - |
| c. Non call rate (0.05, autosome) | 2143/1912 | 675308 | 103 | - |
| d. Sex discrepancy | 2142/1905 | 675308 | 8 | - |
| e. Heterozygosity (-3SD) | 2129/1864 | 675308 | 54 | - |
| **Phase 3: QC, relatedness** | | 675308 | | |
| a. Check for Family IDs | 2129/1864 | 675308 | 0 | - |
| b. Remove close relatives (pihat > 0.2 ) | 2102/1826 | 675308 | 65 | - |
| **Phase 4: QC on SNPs** | | | | |
| a. Remove ChrY | 2102/1826 | 671902 | - | 3406 |
| b. Non call rate (0.05) | 2102/1826 | 666314 | - | 5588 |
| c. +Minor allele freq (0.01) | 2102/1826 | 509663 | - | 156651 |
| d. +Hardy-Weinberg equilibrium (0.00125) | 2102/1826 | 505979 | - | 3684 |
| **Phase 5: Check against 1000G** (McCarthy tool) | | | | |
| a. No Match to 1000G | 2102/1826 | | - | 191 |
| b. Removed for allele freq diff > 0.2 | 2102/1826 | | - | 817 |
| c. Palindromic SNPs with freq > 0.4 | 2102/1826 | | - | 0 |
| d. Non Matching alleles | 2102/1826 | | - | 389 |
| e. Duplicates removed | 2102/1826 | 504056 | - | 526 |

+ Based on European ancestry.

**Table S2.** Details of QC for LifeGene-ANGI-Wave-1.

| | Individuals | SNPs | Removed individuals in each step | Removed SNPs in each step |
|---|---|---|---|---|
| Samples | 1500 | 688032 | - | - |
| **Phase 1: Pre-QC** | | | | |
| a. Check duplicate marker names | 1500 | 688032 | - | 0 |
| b. SNPs not containing rs as part of the name | 1500 | 650645 | - | 37387 |
| c. Remove SNPs without location | 1500 | 650645 | - | 0 |
| d. Remove SNPs on PAR and MT | 1500 | 650641 | - | 4 |
| e. Remove all homozygous SNPs | 1500 | 650641 | - | 0 |
| f. INDELs | 1500 | 650641 | - | 0 |
| g. Remove SNPs sharing the same location | 1500 | 650641 | - | 0 |
| h. Remove ambiguous SNPs | 1500 | 642436 | - | 8205 |
| i. Non call rate on SNPs (0.15) | 1500 | 637487 | - | 4949 |
| **Phase 2: QC on individuals** | | | | |
| a. Check for duplicate samples IDs | 1500 | 637487 | 0 | - |
| b. Remove samples with plating issues | 1500 | 637487 | 0 | - |
| c. Non call rate (0.05, autosome) | 1500 | 637487 | 0 | - |
| d. Sex discrepancy | 1496 | 637487 | 4 | - |
| e. Heterozygosity (-3SD) | 1496 | 637487 | 12 | - |
| **Phase 3: QC, relatedness** | | | | |
| a. Check for Family IDs | 1496 | 637487 | 0 | - |
| b. Remove close relatives (pihat > 0.2 ) | 1454 | 637487 | 30 | - |
| **Phase 4: QC on SNPs** | | | | |
| a. Remove ChrY | 1454 | 637487 | - | 0 |
| b. Non call rate (0.05) | 1454 | 631352 | - | 6135 |
| c. +Minor allele freq (0.01) | 1454 | 491921 | - | 139431 |
| d. +Hardy-Weinberg equilibrium (0.00125) | 1454 | 487997 | - | 3924 |
| a. No Match to 1000G | 1454 | | | |
| b. Removed for allele freq diff > 0.2 | 1454 | | | |
| c. Palindromic SNPs with freq > 0.4 | 1454 | | | |
| d. Non Matching alleles | 1454 | | | |
| e. Duplicates removed | 1454 | 476118 | | |
| f. ** Harmonize to the cases (EGOS) | 1454 | 464330 | | 11798 |

+ Based on European ancestry.

**Genotype Harmonizer software was used for strand alignment and format conversion for genotype data integration between different cohorts/batches. We harmonized all cohorts against the EGOS cohort.

**Table S3.** Details of QC for LifeGene-ANGI-Wave-2.

| | Individuals | SNPs | Removed individuals in each step | Removed SNPs in each step |
|---|---|---|---|---|
| Samples | 1500 | 688032 | - | - |
| **Phase 1: Pre-QC** | | | | |
| a. Check duplicate marker names | 1500 | 688032 | - | 0 |
| b. SNPs not containing rs as part of the name | 1500 | 650645 | - | 37387 |
| c. Remove SNPs without location | 1500 | 650641 | - | 0 |
| d. Remove SNPs on PAR and MT | 1500 | 650641 | - | 4 |
| e. Remove all homozygous SNPs | 1500 | 650641 | - | 0 |
| f. INDELs | 1500 | 650641 | - | 0 |
| g. Remove SNPs sharing the same location | 1500 | 650641 | - | 0 |
| h. Remove ambiguous SNPs | 1500 | 642436 | - | 8205 |
| i. Non call rate on SNPs (0.15) | 1500 | 638254 | - | 4182 |
| **Phase 2: QC on individuals** | | | | |
| a. Check for duplicate samples IDs | 1500 | 638254 | 0 | - |
| b. Remove samples with plating issues | 1500 | 638254 | 0 | - |
| c. Non call rate (0.05, autosome) | 1500 | 638254 | 0 | - |
| d. Sex discrepancy | 1497 | 638254 | 3 | - |
| e. Heterozygosity (-3SD) | 1479 | 638254 | 18 | - |
| **Phase 3: QC, relatedness** | | | | |
| a. Check for Family IDs | 1479 | 638254 | 0 | - |
| b. Remove close relatives (pihat > 0.2 ) | 1458 | 638254 | 21 | - |
| **Phase 4: QC on SNPs** | | | | |
| a. Remove ChrY | 1458 | 638254 | - | 0 |
| b. Non call rate (0.05) | 1458 | 632688 | - | 5566 |
| c. [+]Minor allele freq (0.01) | 1458 | 489692 | - | 142996 |
| d. [+]Hardy-Weinberg equilibrium (0.00125) | 1458 | 487931 | - | 1761 |
| a. No Match to 1000G | 1458 | | - | |
| b. Removed for allele freq diff > 0.2 | 1458 | | - | |
| c. Palindromic SNPs with freq > 0.4 | 1458 | | - | |
| d. Non Matching alleles | 1458 | | - | |
| e. Duplicates removed | 1458 | 475940 | - | |
| f. [**]Harmonize to the cases (EGOS) | 1458 | 465124 | - | 10816 |

[+] Based on European ancestry (the largest clusters in GEMToolss).
** Genotype Harmonizer software was used for strand alignment and format conversion for genotype data integration between different cohorts/batches. We harmonized all cohorts against the EGOS cohort.

### Population stratification, ancestry groups

We used GEMTools to find individuals with recent European ancestry. GEMTools uses spectral graph methods to find a low-dimensional representation of the genetic similarities between individuals, which is referred to as an eigenmap. Assuming an eigenmap is constructed using a representative base sample, additional individuals can be projected onto the map using the Nystrom approximation (1). Non-base individuals are assigned to the cluster of their genetically closest base-neighbor.

Figure S1 illustrates the base and non-base individuals for the first six ancestry vectors. Individuals in clusters A, B, C, and D have the closest ancestry (min.dim=6; GEMTools found two eigenvectors without using min.dim).

**Figure S1.** Results from GEMTools (colors represent the base and non-base individuals).

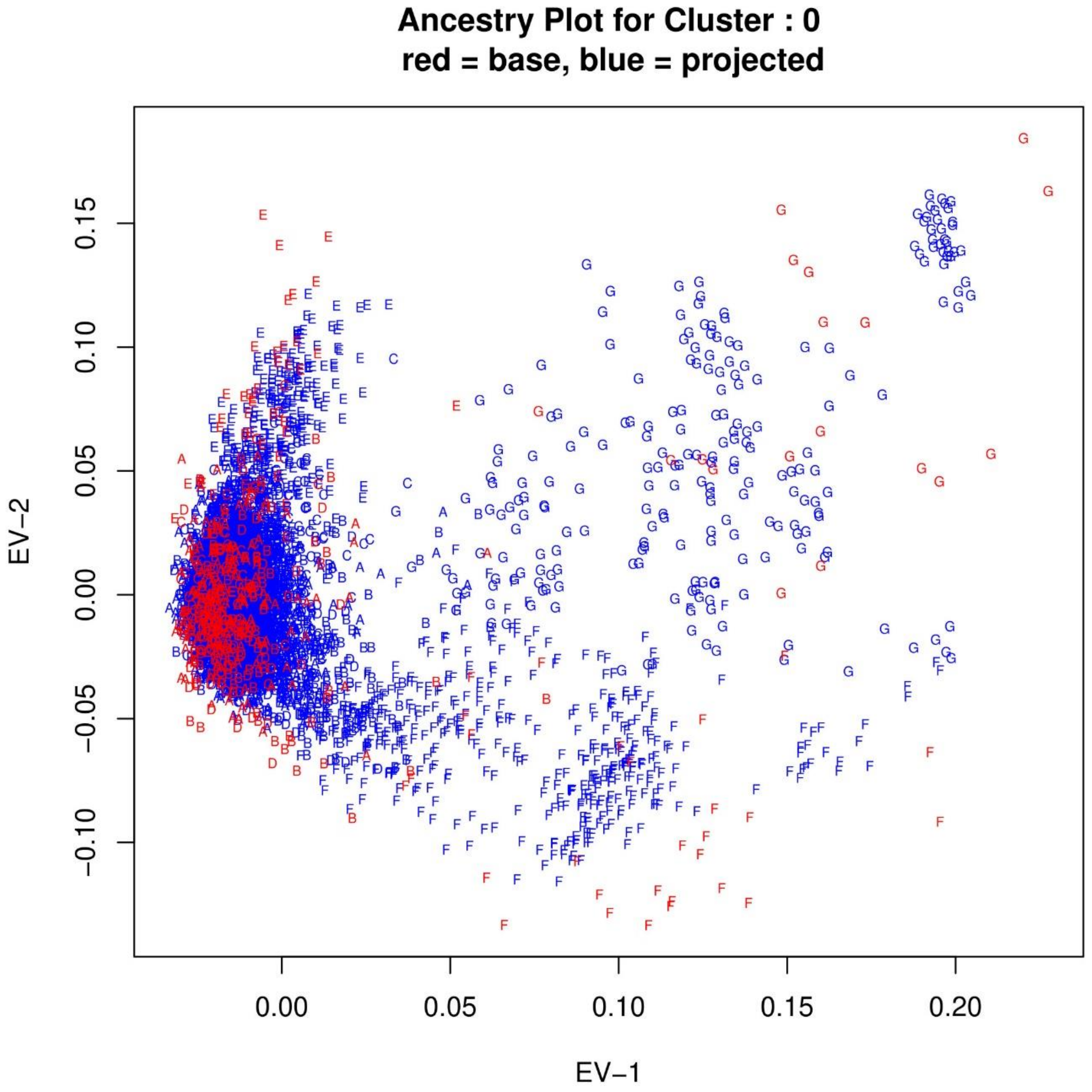

### Principle component analysis (PCA) for population structure

We used PLINK 2.0 to calculate the first 20 PCAs (after linkage disequilibrium (LD) pruning of the SNPs, --indep 50 5 0.2). The first six PCAs explained around 70% of the variance discovered by the first 20 PCAs (Figure S2). Therefore, we used the first six PCAs to adjust for population structure.

**Figure S2** The ratio of each eigenvalue to the sum of PCAs.

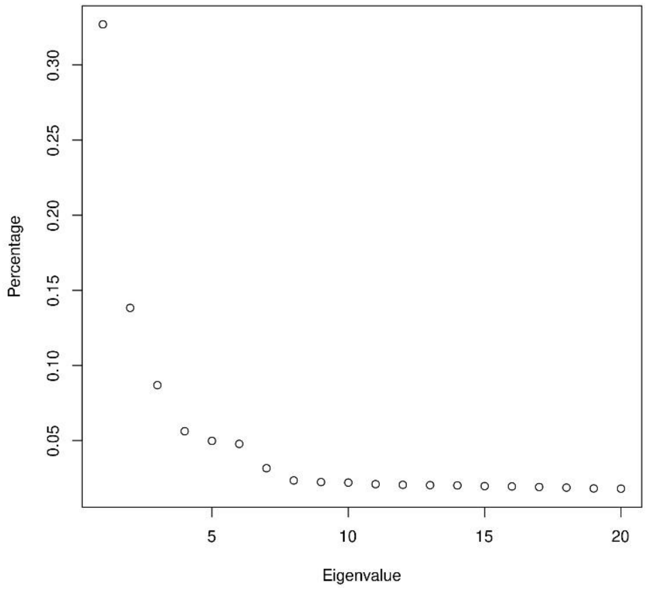

### Heritability for different population prevalences

Table S4 shows the estimate of heritability for different population prevalences (using the first 6 PCAs as covariates). The source population for EGOS is from the Swedish National Patient Register (NPR) and most of the NORDiC cases can be found in NPR. Previously, we estimated 0.0087 as the population prevalence of OCD for individuals born in Sweden between 1982-1990 and have a diagnosis in NPR (2).

**Table S4.** Estimates of heritability for different population prevalence

| Prevalence | heritability (SE) |
|---|---|
| 0.005 | 24% (3%) |
| 0.01 | 28% (4%) |
| 0.015 | 31% (5%) |
| 0.02 | 34% (5%) |
| 0.025 | 36% (5%) |
| 0.03 | 37% (5%) |

### Comparison of EGOS and NORDiC cases

Principle component analysis of the first two ancestry vectors for cases and controls are illustrated in Figure S3. For illustration purposes, we focused on individuals with PCA-1 < 0 (Figures S3).

**Figure S3.** First two ancestry vectors.

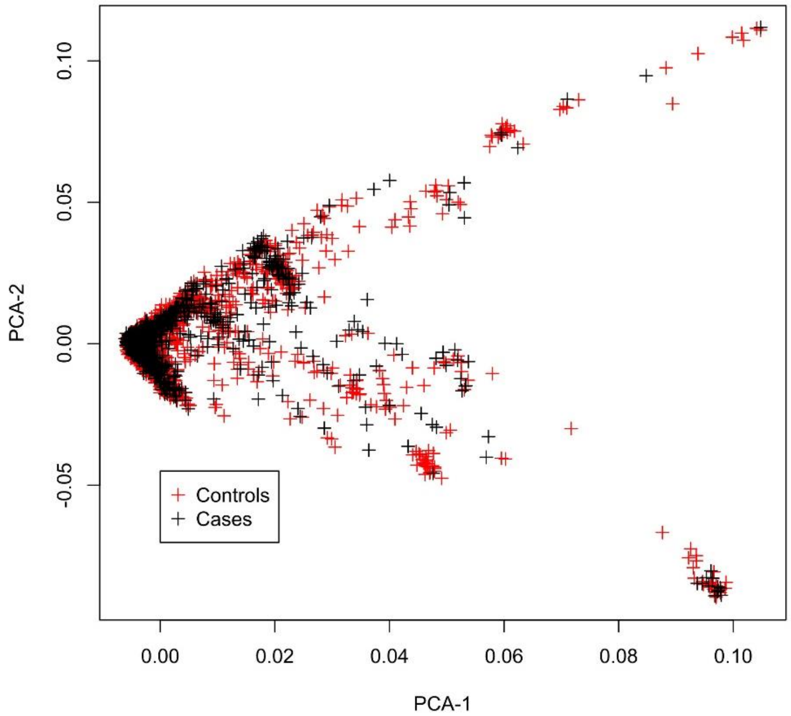

Figures S4 and S5 show the PCAs for the controls and cases, respectively (for PCA-1 < 0). Figures S6 and S7 show the PCAs for EGOS and NORDiC cases.

**Figure S4.** Controls, the first two ancestry vectors.

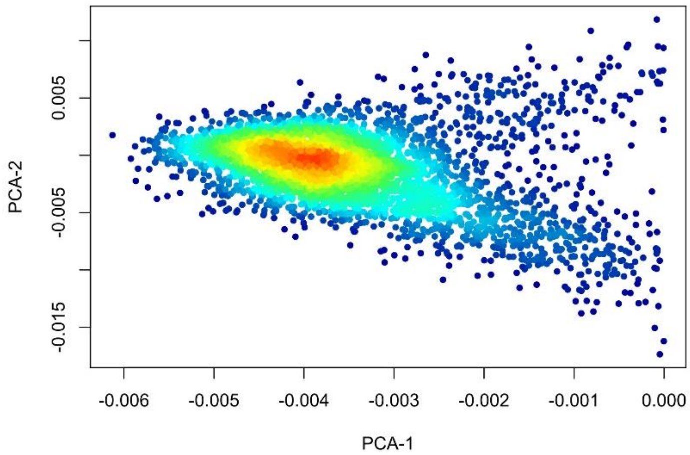

**Figure S5.** All cases, the first two ancestry vectors.

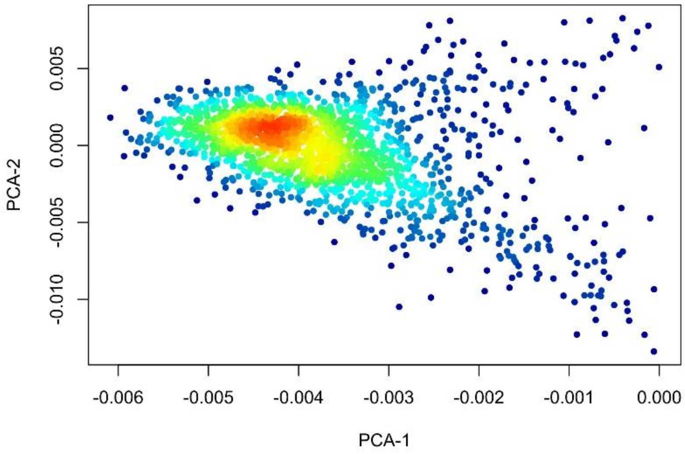

**Figure S6.** EGOS, the first two ancestry vectors.

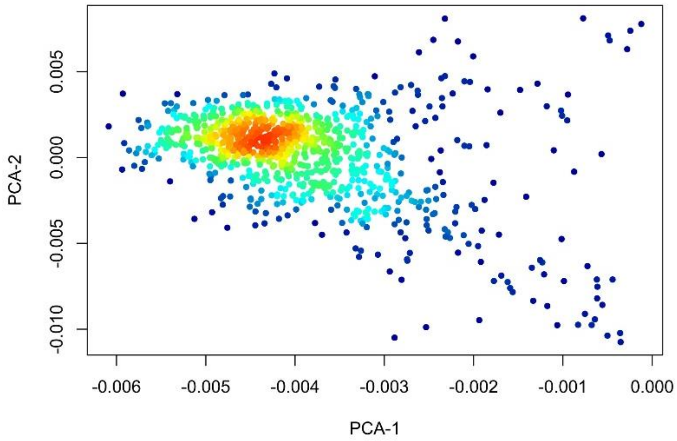

**Figure S7.** NORDiC, the first two ancestry vectors.

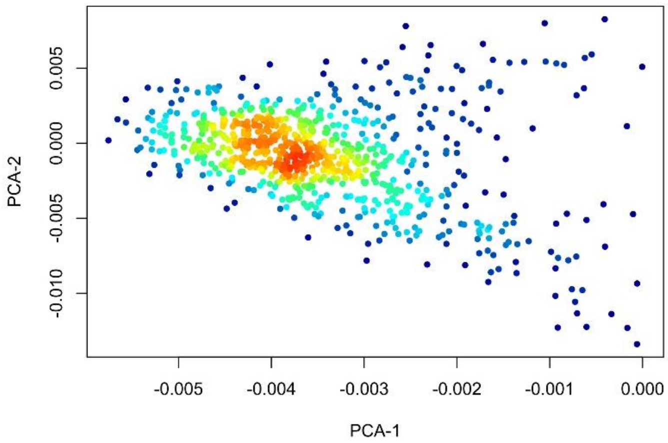

Comparison of Figures S6 and S7 suggests that EGOS and NORDiC cases have slightly different ancestry distribution. EGOS cases are more concentrated above zero for PCA-2. We observed a similar pattern in the histograms of PCA-2 in Figures S8 and S9. The ancestry distribution of EGOS cases was not a perfect match to that of controls. However, when EGOS and NORDiC were merged, their ancestry distribution matched the controls quite well (Figure S4 and S5).

**Figure S8.** EGOS cases, first two ancestry vectors.

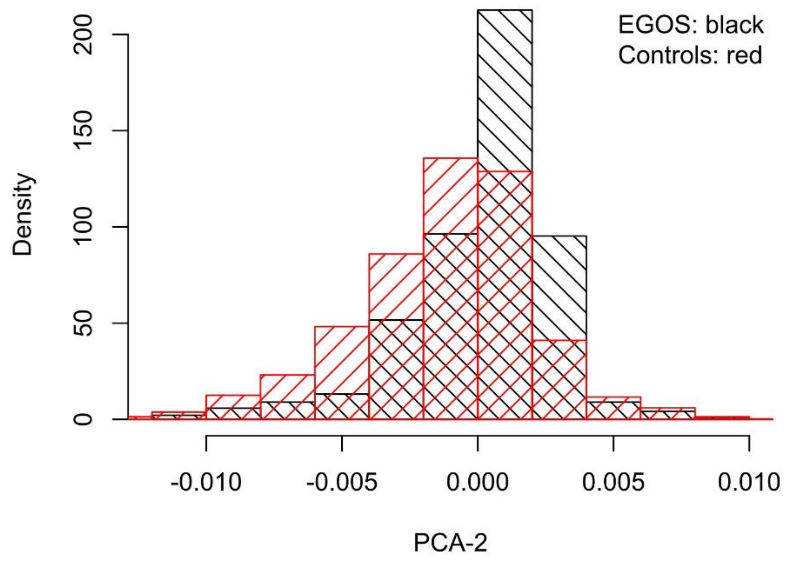

**Figure S9.** NORDiC cases, first two ancestry vectors.

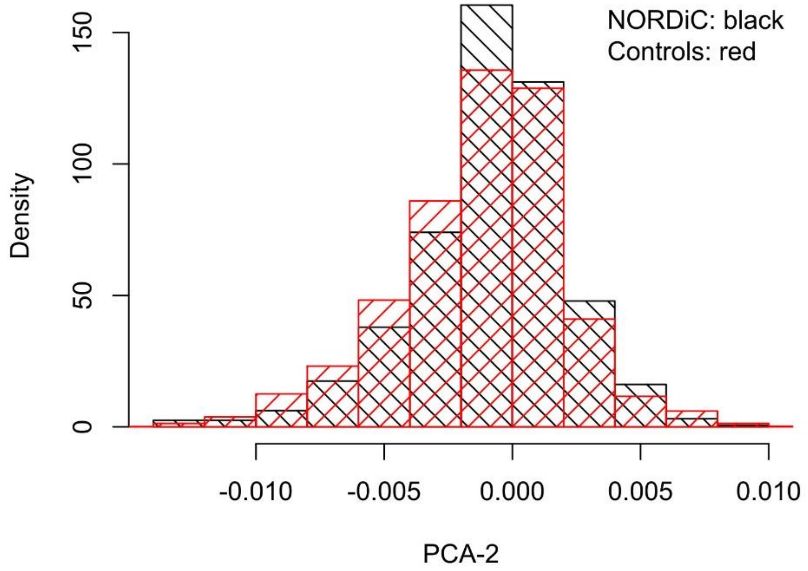

We used 1:1 pair matching using PCA-1 and PCA-2 as the distance function (*pairmatch* function in R). EGOS and NORDiC cases had similar heritability after matching controls (Table S5).

**Table S5.** Estimates of heritability for EGOS and NORDiC cases.

| | Heritability (SE) |
|---|---|
| EGOS and matched controls | 28% (11%) |
| NORDiC and matched controls | 27% (11%) |

### Heritability analysis partitioned by MAF bins

**Table S6.** Heritability estimates for ten samples of size 180K SNPs. Sampling from each bin was proportional to the percentage of SNPs in that bin in the real data.

| MAF | SNPs | % of the total SNPs | Heritability (10 Samples) | | | | | | | | | | | % of heritability |
|---|---|---|---|---|---|---|---|---|---|---|---|---|---|---|
| | | | 1 | 2 | 3 | 4 | 5 | 6 | 7 | 8 | 9 | 10 | Mean | |
| **0.01-0.05** | 82080 | 45.6% | 2.4 | 3.7 | 4.8 | 6.2 | 1.1 | 1.6 | 3.6 | 3.4 | 3.1 | 3.5 | 3.3 | 15.3% |
| **0.05-0.1** | 21420 | 11.9% | 1.7 | 0.0 | 0.0 | 1.6 | 1.4 | 1.1 | 0.0 | 0.0 | 0.0 | 0.5 | 0.6 | 2.8% |
| **0.1-0.2** | 25740 | 14.3% | 3.2 | 2.3 | 2.5 | 1.8 | 3.1 | 1.6 | 3.5 | 2.3 | 2.8 | 4.0 | 2.7 | 12.5% |
| **0.2-0.3** | 19800 | 11.0% | 5.7 | 6.6 | 5.4 | 5.1 | 7.9 | 6.7 | 5.7 | 5.5 | 7.0 | 6.6 | 6.2 | 28.7% |
| **0.3-0.4** | 16020 | 8.9% | 4.1 | 3.2 | 3.8 | 2.7 | 2.9 | 4.6 | 3.5 | 1.8 | 3.9 | 2.3 | 3.3 | 15.3% |
| **0.4-0.5** | 14940 | 8.3% | 3.2 | 5.9 | 5.7 | 6.4 | 5.3 | 6.3 | 4.1 | 6.9 | 5.0 | 5.8 | 5.5 | 25.5% |
| **Total** | 180000 | 100% | 20.3 | 21.7 | 22.2 | 23.8 | 21.7 | 21.9 | 20.4 | 19.9 | 21.8 | 22.7 | | |

**Table S7.** Heritability estimates for ten samples of size 180K SNPs. Sampling from each bin was proportional to the percentage of SNPs in that bin from 1000G data.

| MAF | SNPs | % of the total SNPs | Heritability (10 Samples) | | | | | | | | | | | % of heritability |
|---|---|---|---|---|---|---|---|---|---|---|---|---|---|---|
| | | | 1 | 2 | 3 | 4 | 5 | 6 | 7 | 8 | 9 | 10 | Mean | |
| **0.01-0.05** | 53100 | 29.5% | 3.9 | 1.5 | 3.1 | 1.6 | 1.5 | 1.5 | 1.4 | 1.2 | 1.0 | 2.5 | 1.9 | 8.3% |
| **0.05-0.1** | 25200 | 14.0% | 0.0 | 0.0 | 0.7 | 2.0 | 1.1 | 0.0 | 0.9 | 0.6 | 0.3 | 5.5 | 1.1 | 4.8% |
| **0.1-0.2** | 32940 | 18.3% | 4.0 | 5.7 | 3.1 | 2.2 | 3.3 | 3.0 | 4.2 | 1.9 | 5.5 | 2.5 | 3.5 | 15.4% |
| **0.2-0.3** | 25200 | 14.0% | 4.6 | 6.8 | 7.7 | 6.2 | 7.2 | 5.4 | 6.5 | 7.9 | 6.3 | 6.5 | 6.5 | 28.5% |
| **0.3-0.4** | 22320 | 12.4% | 3.8 | 3.1 | 2.9 | 2.9 | 2.3 | 5.1 | 3.5 | 2.4 | 2.5 | 4.2 | 3.3 | 14.5% |
| **0.4-0.5** | 21240 | 11.8% | 6.6 | 5.2 | 6.2 | 6.6 | 7.0 | 6.8 | 7.5 | 6.2 | 5.8 | 7.2 | 6.5 | 28.5% |
| **Total** | 180000 | 100% | 22.9 | 22.3 | 23.7 | 21.5 | 22.4 | 21.8 | 24 | 20.2 | 21.4 | 28.4 | | |

**Table S8.** Heritability estimates for ten samples of size 180K SNPs. 30K samples from each bin.

| MAF | SNPs | % of the total SNPs | Heritability (10 Samples) | | | | | | | | | | | % of heritability |
|---|---|---|---|---|---|---|---|---|---|---|---|---|---|---|
| | | | 1 | 2 | 3 | 4 | 5 | 6 | 7 | 8 | 9 | 10 | Mean | |
| **0.01-0.05** | 30000 | 16.7% | 1.6 | 0.2 | 0.0 | 0.3 | 1.1 | 0.0 | 0.0 | 3.1 | 1.5 | 2.4 | 1.0 | 4.4% |
| **0.05-0.1** | 30000 | 16.7% | 1.2 | 2.4 | 2.0 | 0.0 | 1.2 | 1.8 | 0.1 | 0.0 | 0.6 | 0.9 | 1.0 | 4.4% |
| **0.1-0.2** | 30000 | 16.7% | 1.9 | 1.8 | 2.8 | 3.5 | 1.7 | 2.8 | 3.6 | 2.9 | 3.0 | 1.5 | 2.5 | 11.0% |
| **0.2-0.3** | 30000 | 16.7% | 6.4 | 8.6 | 8.7 | 9.5 | 6.8 | 8.2 | 9.5 | 8.4 | 7.2 | 7.1 | 8.0 | 35.1% |
| **0.3-0.4** | 30000 | 16.7% | 5.3 | 2.7 | 2.4 | 2.6 | 5.1 | 3.2 | 2.9 | 3.1 | 3.9 | 4.6 | 3.6 | 15.8% |
| **0.4-0.5** | 30000 | 16.7% | 7.0 | 7.2 | 6.0 | 6.7 | 6.7 | 6.4 | 6.4 | 6.7 | 6.8 | 6.9 | 6.7 | 29.4% |
| **Total** | 180000 | 100% | 23.4 | 22.9 | 21.9 | 22.6 | 22.6 | 22.4 | 22.5 | 24.2 | 23 | 23.4 | | |

**Figure S10.** The proportion of expected and observed heritability explained by different minor allele frequencies (MAF) bins based on A) the real data; B) the average of ten samples of size 180K SNPs, sampling from each bin was proportional to the percentage of SNPs in that bin in the real data; C) the average of ten samples of size 180K SNPs, sampling from each bin was proportional to the percentage of SNPs in 1000 Genomes data; D) the average of ten samples of size 180K SNPs, 30K samples from each bin. MAFs were binned, and we used the average MAF in a bin to plot the results.

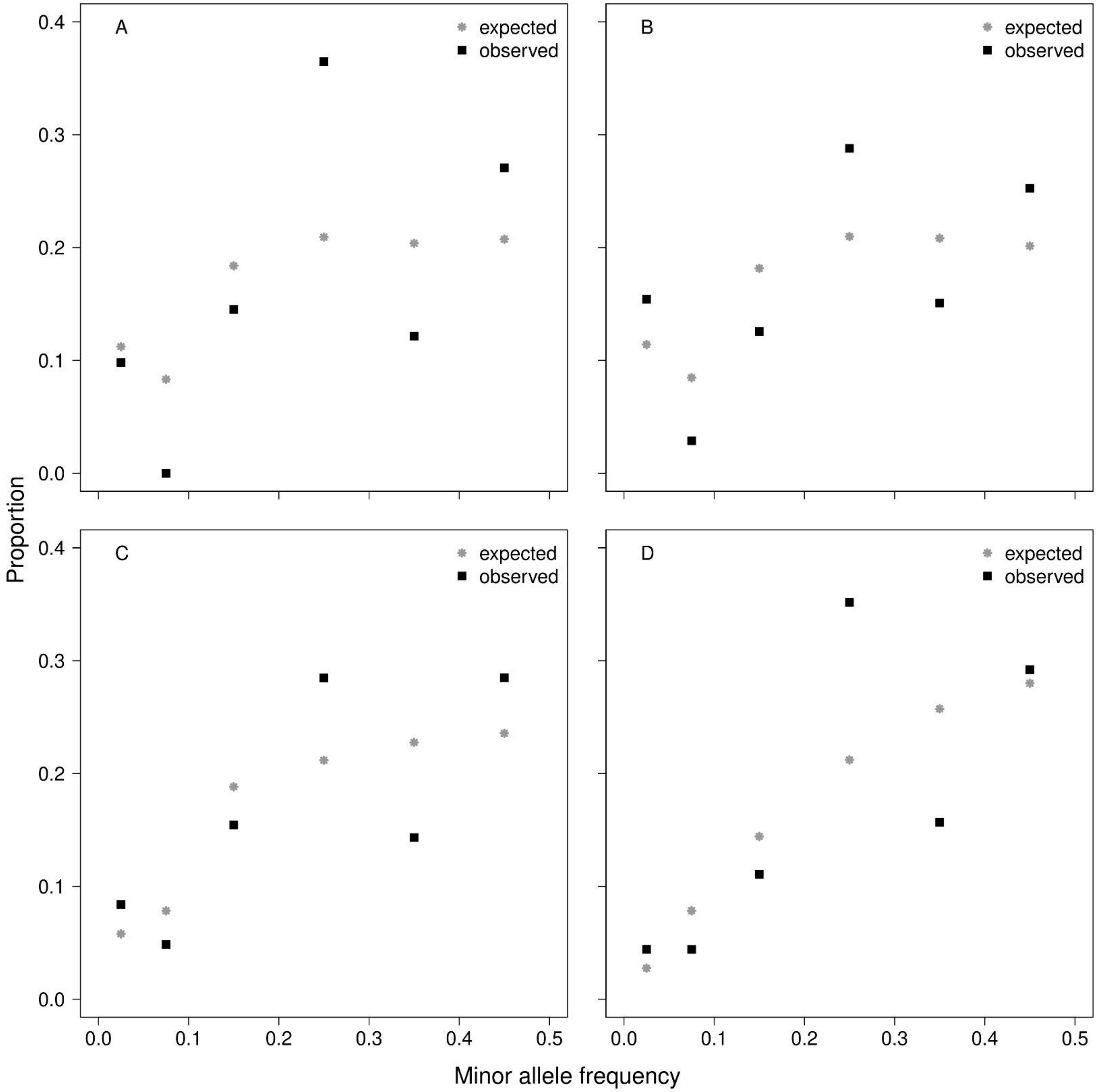

**Figure S11.** The observed proportion of heritability versus its expected proportion based on A) the real data ($R^2$=0.53, p-value=0.062); B) the average of ten samples of size 180K SNPs, sampling from each bin was proportional to the percentage of SNPs in that bin in the real data ($R^2$=0.46, p-value=0.082); C) the average of ten samples of size 180K SNPs, sampling from each bin was proportional to the percentage of SNPs in 1000G data ($R^2$=0.56, p-value=0.054); D) the average of ten samples of size 180K SNPs, 30K samples from each bin. In each plot, the solid line is the regressed line and the dashed line has slope one and intercept zero (observed=expected) ($R^2$=0.51, p-value=0.067).

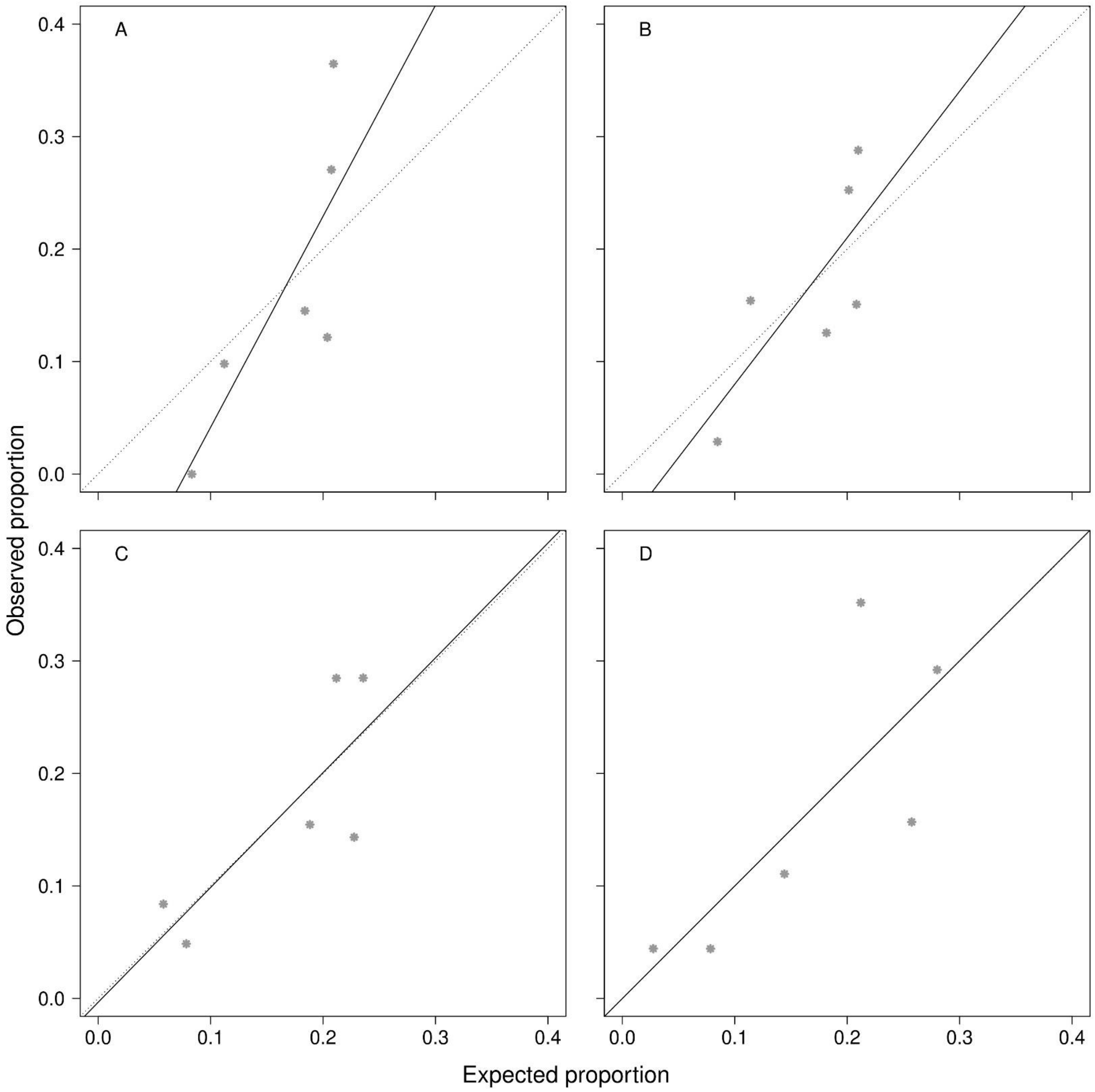

### Analysis of the sex

**Table S9.** Estimates of heritability partitioned by sex

| Cohort | Heritability (SE) Female | Male |
|---|---|---|
| All cases and all controls | 23% (6%) | 9% (12%) |
| All cases, excluding LifeGene-ANGI controls | 0.0001% (13%) | 2 % (11%) |
| All cases, excluding LifeGene-ANGI controls, pair-matched controls | 0.0001% (17%)[1] | 0.0001% (13%) |

[1] There were not enough controls for a full match (588 cases were removed).

**Table S10.** Estimates of heritability partitioned by MAF bins in this study, in Davis *et al.* (3) and proportional to 1000G data. For 1000G proportional to data, the estimate of heritability for each bin is the mean of heritability for that bin for ten samples of size 108K SNP; Sampling from each bin was proportional to the percentage of SNPs in that bin from 1000G data.

| | This manuscript | | | Davis *et al.* (5) | | | Expected (Proportional to 1000G) | | |
|---|---|---|---|---|---|---|---|---|---|
| **MAF** | **Heritability (SE)** | **SNPs (% of total)** | **% Heritability** | **Heritability (SE)** | **SNPs (% of total)** | **% Heritability** | **Heritability (SE)[2]** | **SNPs (% of total)** | **% Heritability** |
| **0.01-0.05** | 2.5% (3.7%) | 181673 (45.6%) | 9.7% | 0.0001% (3%)[1] | 19605 (5.2) | 0% | 1.9% (0.3%) | 53100 (29.5%) | 8.3% |
| **0.05-0.1** | 0.0% (2.0%) | 47404 (11.9%) | 0.0% | 4% (5%) | 47976 (12.8) | 11% | 1.1% (0.5%) | 25200 (14.0%) | 4.8% |
| **0.1-0.2** | 3.7% (2.4%) | 56918 (14.3%) | 14.2% | 8% (6%) | 91661 (24.5) | 23% | 3.5% (0.4%) | 32940 (18.3%) | 15.4% |
| **0.2-0.3** | 9.3% (2.3%) | 44043 (11%) | 36.6% | 1% (6%) | 77193 (20.7) | 3% | 6.5% (0.3%) | 25200 (14.0%) | 28.5% |
| **0.3-0.4** | 3.1% (2.1%) | 35349 (8.9%) | 12.4% | 11% (5%) | 70193 (18.7) | 31% | 3.3% (0.3%) | 22320 (12.4%) | 14.5% |
| **0.4-0.5** | 6.9% (2.0%) | 33069 (8.3%) | 27.1% | 11% (5%) | 66770 (17.8) | 31% | 6.5% (0.2%) | 21240 (11.8%) | 28.5% |
| Sum | 25.5% | 398456 | 100% | 35% | 373398 | 100% | 22.8% | 180000 | 100% |

[1]The reported boundary for this study was 0.001-0.05. [2]The estimated standard error based on the ten samples.
